# Longitudinal intronic RNA-Seq analysis of Parkinson’s Disease patients reveals disease-specific nascent transcription

**DOI:** 10.1101/2021.11.03.21265851

**Authors:** Sulev Kõks, Abigail L. Pfaff, Vivien J. Bubb, John P. Quinn

**Affiliations:** Perron Institute for Neurological and Translational Science, Perth, WA 6009, Australia; Centre for Molecular Medicine and Innovative Therapeutics, Murdoch University, Perth, WA 6150, Australia; Department of Pharmacology and Therapeutics, Institute of Systems, Molecular and Integrative Biology, University of Liverpool, Liverpool L69 3BX, UK

**Keywords:** Parkinson Disease, transcriptome, whole transcriptome analysis, introns, RNA-Seq, nascent transcript, nascent RNA, PPMI, blood transcriptome

## Abstract

Transcriptomic studies usually focus on either gene or exon-based annotations, and only limited experiments have reported changes in reads mapping to introns. The analysis of intronic reads allows the detection of nascent transcription that is not influenced by steady - state RNA levels and provides information on actively transcribed genes. Here we describe substantial intronic transcriptional changes in Parkinson’s Disease (PD) patients compared to healthy controls (CO) at two different timepoints; at the time of diagnosis (BL) and three years later (V08). We used blood RNA-Seq data from the Parkinson’s Progression Markers Initiative (PPMI) cohort and identified significantly changed transcription of intronic reads only in PD patients during this follow up period. In CO subjects, only nine transcripts demonstrated differentially expressed introns between visits. However, in PD patients 4,873 transcripts had differentially expressed introns at visit V08 compared to BL, many of them in genes previously associated with neurodegenerative diseases, such as *LRRK2, C9orf72, LGALS3, KANSL1AS1* and *ALS2*. In addition, at the time of diagnosis (BL visit) we identified 836 transcripts (e.g. *SNCA, DNAJC19, PRRG4*) and at visit V08 2,184 transcripts (e.g. *PINK1, GBA, ALS2, PLEKHM1*) with differential intronic expression specific to PD patients. In contrast, reads mapping to exonic regions demonstrated little variation indicating highly specific changes only in intronic transcription. Our study demonstrated that Parkinson’s disease is characterized by substantial changes in the nascent transcription and description of these changes could help to understand the molecular pathology underpinning this disease.

**Impact statement:** Transcriptomic studies in most cases describe the steady state changes of the cellular RNA combined with signals from newly synthesised RNA or nascent RNA. Nascent RNA reflects dynamic alterations in the cellular transcriptome and improves the resolution of RNA-Seq analysis. In the present study, we describe the changes in nascent RNA transcription in Parkinson’s disease by using intronic RNA-Seq analysis. We compared transcriptome changes at the time of diagnosis and 3 years after the initial diagnosis. As a result, we were able to describe disease-specific time-dependent alterations in the nascent transcription in the blood of Parkinson’s patients illustrating another layer of the blood-based biomarkers that could be diagnostic of both risk and progression of Parkinson’s disease.

## Introduction

The analysis of the transcriptome is usually based on reads generated from exons and gene-based annotations. A large proportion of these reads from RNA-sequencing map to the intronic sequences and this is true for both ribosomal depleted and poly-A selected RNA protocols ^1^. It has been described that 38% of total RNA-Seq reads and only 8% of polyA RNA reads map to introns ^1^. The significance of the intronic reads has remained controversial and this part of the transcriptome is mostly neglected in transcriptomic analyses ^2^. However, recent studies have shown that reads mapping to introns reflect the immediate regulatory responses in transcription compared to post-transcriptional or steady-state changes ^2, 3^. The intronic reads reflect the presence of the newly transcribed RNA and as such are useful to explore the complexity of nascent RNA transcription and co-transcriptional splicing ^4-6^. The analysis of intronic reads has been used to develop a detailed transcriptional model within a single sample showing the utility of intronic reads to estimate the genome-wide pre-mRNA synthesis rate ^7^, and that intronic coverage was related to nascent transcription and co-transcriptional splicing ^1^. The levels of intronic reads can precede the change in exonic reads by 15 minutes, making them very useful to detect immediate responsive changes in the transcriptome ^5^. Therefore, analysis of the intronic reads allows the separation of nascent transcription from post-transcriptional changes during the formation of the genome-wide transcriptome.

Parkinson’s Disease (PD) is one of the most common neurodegenerative diseases with several genetic mutations and variants associated with the disease ^8-11^. The exact mechanisms underpinning risk and progression of the disease are still not clear and its neuropathology complex. Therefore, the genomic network leading to pathology is also likely to be both complex and multifactorial ^12-14^. Several studies have performed transcriptomic analysis in PD identifying transcriptional signatures specific to the disease or involved in the regulation of splicing of genes expressed in the basal ganglia ^15, 16^. The analysis of peripheral tissue transcriptomes (blood and skin) overlaid on CNS PD RNA-Seq data has identified significant differences between PD and control transcriptomic profiles ^12, 17, 18^. Those studies underlined the importance of peripheral tissue analysis to both determine biomarkers correlating with the disease and gain insight into signalling pathways operating in neurodegenerative diseases. In this longitudinal study utilising data from the Parkinson’s Progression Markers Initiative (PPMI) cohort we compared the blood intronic transcriptome of PD patients and healthy controls (Table 1) at two different timepoints of disease progression, at diagnosis (baseline, BL) and after three years follow up (V08). This allowed us to identify the immediate changes in the transcriptome reflected by the changes in nascent RNA using only reads mapping to the intronic sequences. Intronic data were compared with the exonic reads to differentiate steady-state transcription from nascent transcription.

**Table 1.** Overview of the study samples and design. Subject numbers with the blood whole transcriptome data of the PPMI cohort are given.

| Study group | Baseline (BL) | Three years (V08) |
| --- | --- | --- |
| CO | 189 | 157 |
| PD | 390 | 338 |

## Materials and Methods

### Datasets

In this study we utilized the Parkinson’s Progression Markers Initiative (PPMI) cohort data that were downloaded from www.ppmi-info.org/data (16 May 2021). The PPMI cohort of Parkinson’s patients containing longitudinal data with the aims to describe the progression and biomarkers of Parkinson’s disease (PD). It is an observational, multi-centre natural history study for PD. PPMI assesses progression of clinical features, imaging outcomes, biological and genetic markers and digital outcomes of PD across different stages of disease from prodromal to moderate. The overall goal of the study is to identify markers of disease progression to accelerate therapeutic trials to reduce progression of PD disability. The clinical protocol is designed to acquire comprehensive longitudinal within-participant data in approximately 4,000 participants enrolled at approximately 50 sites worldwide.

The PPMI dataset we used in our current study contains whole transcriptome data from the blood together with genetic and clinical data from patients with verified PD and control status. For the RNA-Seq 1μg of RNA isolated from PaxGene tubes was used and sequencing was performed at Hudson’s Alpha’s Genomic Services Lab on an Illumina NovaSeq6000. All samples underwent rRNA and globin reduction, followed by directional cDNA synthesis using the NEB kit. Following second-strand synthesis, library samples were prepared using the NEB/Kapa (NEBKAP) based library kit. Fastq files were merged and aligned to GRCh38p12 by STAR ^19^ (v2.6.1d) on GENCODE v29. The specific options for the STAR alignment workflow were following: --runMode alignReads --twopassMode Basic -- outSAMtype SAM --outFilterType BySJout --outFilterMultimapNmax 20 -- outFilterMismatchNmax 999 --outFilterMismatchNoverLmax 0.1 --alignIntronMax 1000000 --alignMatesGapMax 1000000 --alignSJoverhangMin 8 --alignSJDBoverhangMin 1 --chimSegmentMin 15 --chimJunctionOverhangMin 15 –outSAMstrandField intronMotif -- outSAMunmapped Within --outSAMattrRGline

### Analytical Workflow

Bam files of RNA-Seq data from PPMI were imported to the R environment and intronic reads were called using packages “GenomicFeatures” and “GenomicAlignments” ^20^. Initially we built a genome-wide intron-containing list of features by selecting intronic information from the TxDb object that was based on “gencode.v38.chr_patch_hapl_scaff.annotation.gtf”. To retrieve intronic data from txDb object, “tidyIntrons’ function was used. The function “tidyIntrons” returns a GRanges object with 1 range per intron and with metadata columns tx_id, tx_name, and gene_id. We next applied “summarizeOverlaps” with the “union” mode of counting to get raw expression counts for every intron. Normalization was performed during differential expression analysis by using the median of ratios method. Briefly, counts were divided by sample-specific size factors that were defined as a median ratio of gene counts to the geometric mean per gene. Differential expression was detected using “DESeq2” and functionally annotated using “ReactomePA”, “DOSE” and “clusterProfiler” packages.

### Statistical analysis

Formal statistical analysis of differential expression of the intronic transcripts was performed by using the “DESeq2” packages for R and only False Discovery Rate (FDR) corrected transcripts below 0.05 are reported. In addition, selection of FDR corrected transcripts was used for the pairwise comparison of the expression data and the plots were generated using “ggpubr” package.

## Results

We analysed the intronic expression profiles in PD and CO subjects at the time of diagnosis (BL visit) and three years later (V08). There was little differential expression in exons in all our comparisons in both PD and CO groups. However, there were many changes observed in intronic expression in the PD cohort which were highly significant and consistent with widespread active transcription in the blood from BL to V08. In addition, only a few active intronic transcriptional changes were observed in the CO subjects at BL and V08 timepoints (Table 2). In contrast, very few differences in transcripts were identified in exons (Table 2, Figure 1). More detailed description of our findings follows.

**Table 2.** Comparison of the differential expression of the intronic and exonic reads. For baseline (BL) and three-year (V08) time points PD was compared to CO. For PD and CO groups, the effect of time (three years) was measured (V08 versus BL), significance threshold was FDR < 0.05.

| Group | Introns | Exons | Comparison |
| --- | --- | --- | --- |
| BL | 836 | 1 | PD versus CO |
| V08 | 2184 | 17 | PD versus CO |
| PD | 4873 | 8 | V08 versus BL |
| CO | 9 | 9 | V08 versus BL |

**Figure 1.**
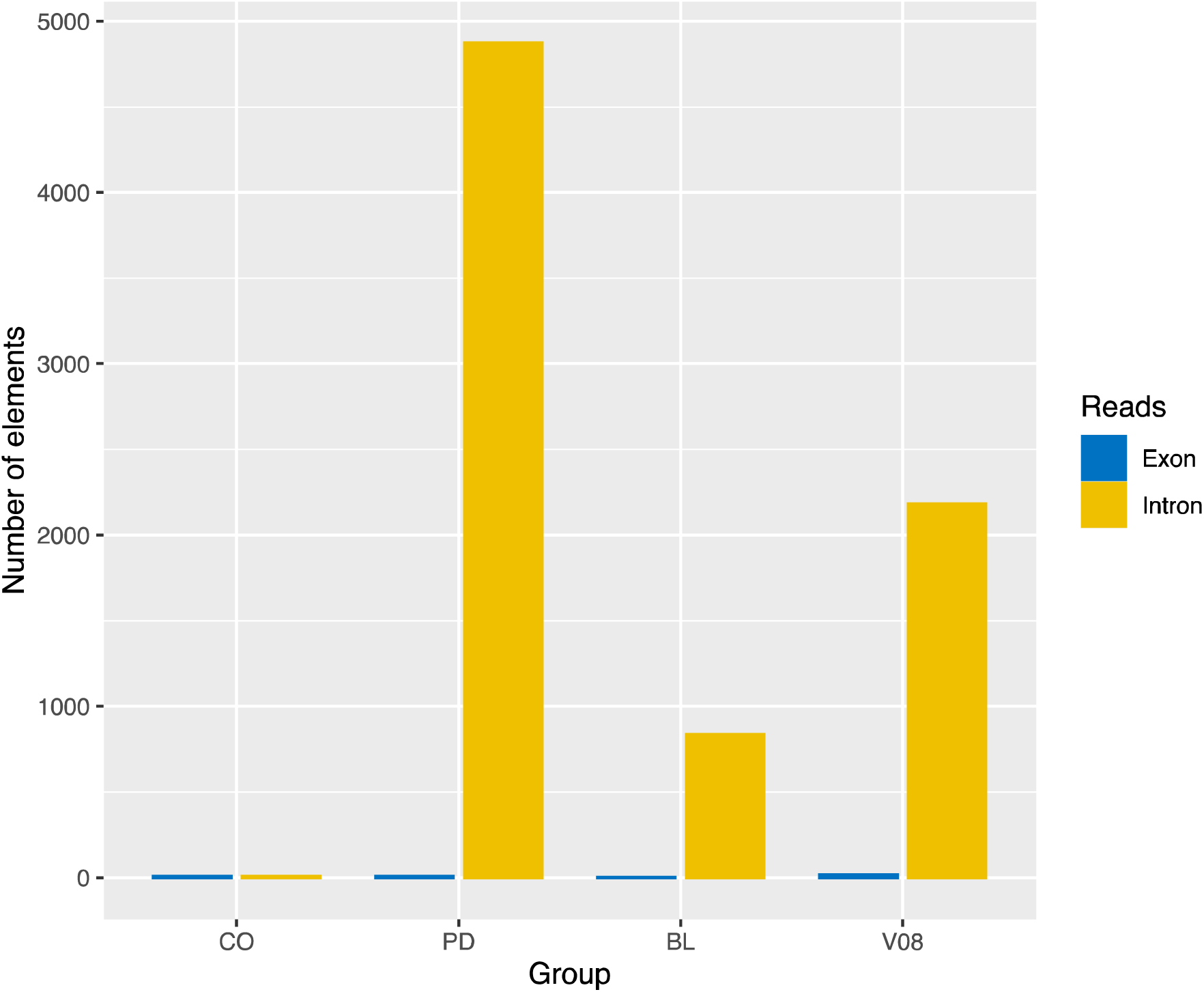
The figure illustrates the number of differentially expressed exonic and intronic reads in different subgroups and at different timepoints. The number of intronic reads is very high in PD patients compared to CO subjects. Exonic reads didn’t change by the disease status or by the progression of the disease. It is also evident that the number of intronic reads increases from the baseline visit to three-year follow up.

### CO subjects

We analysed the longitudinal changes in the expression of the intronic reads of control subjects at BL and V08 timepoints and identified only limited differences. Nine transcripts showed differential intronic expression below FDR 0.05 (Supplementary Table 1). We also analysed the differential expression of the exonic transcripts, and nine transcripts were differentially expressed with exonic reads (Supplementary Table 2). This demonstrates that differential expression in control subjects is quite limited, in both the intronic and exonic analysis, and we can conclude that the transcriptome for control subjects is longitudinally very stable.

### PD subjects

We analysed PD patients and compared their longitudinal intronic and exonic transcriptional profile. We identified 4,873 differentially expressed introns in PD patients that differed between the time of diagnosis and after three years follow up (Supplementary Table 3). These 4,873 introns reflect the longitudinal change that is specific for PD patients during the three-year period. The CO cohort in comparison exhibited a very stable transcriptome with very limited changes. In contrast to intronic changes we detected only 8 exonic reads which were differentially expressed, (Supplementary Table 4). Within the differentially expressed introns we found many that were in genes known to be involved in neurodegenerative diseases and PD, such as *LRRK2, VPS13C, LGALS3, C9orf72* and *ALS2* (Figure 2). We also identified statistically significant upregulation of intron 1 and 2 expression of the PD associated gene PINK1 gene, showing that the intronic changes are not limited to single introns (Figure 3). *FLACC1, KANLS1AS1* and *CASP8AP2* were other genes in which differential intron expression occurred (Figure 3). Taken together, we can conclude here that PD patients express overwhelming longitudinal changes in nascent transcription.

**Figure 2.**
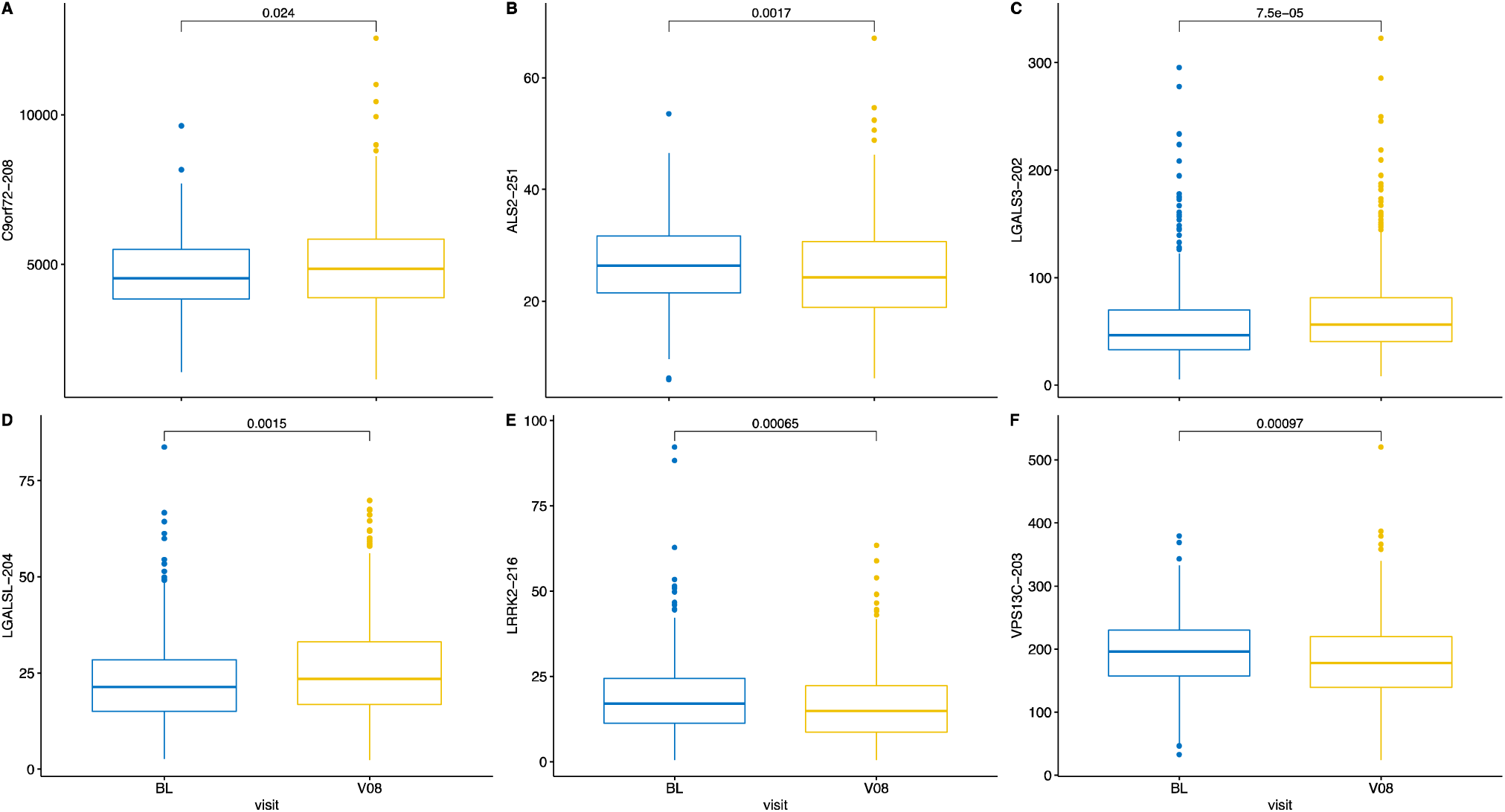
Longitudinal differential expression (normalised counts) of the intronic transcripts in PD patients, baseline (BL) compared to the V08 visit three years later. Panels A, B, C, D, E, F reflect changes in the introns for different transcripts. LRRK2, ALS2 and VPS13C are downregulated during the progression of PD. C9orf72, LGALS3 and LGALSL are upregulated within three years of PD.

**Figure 3.**
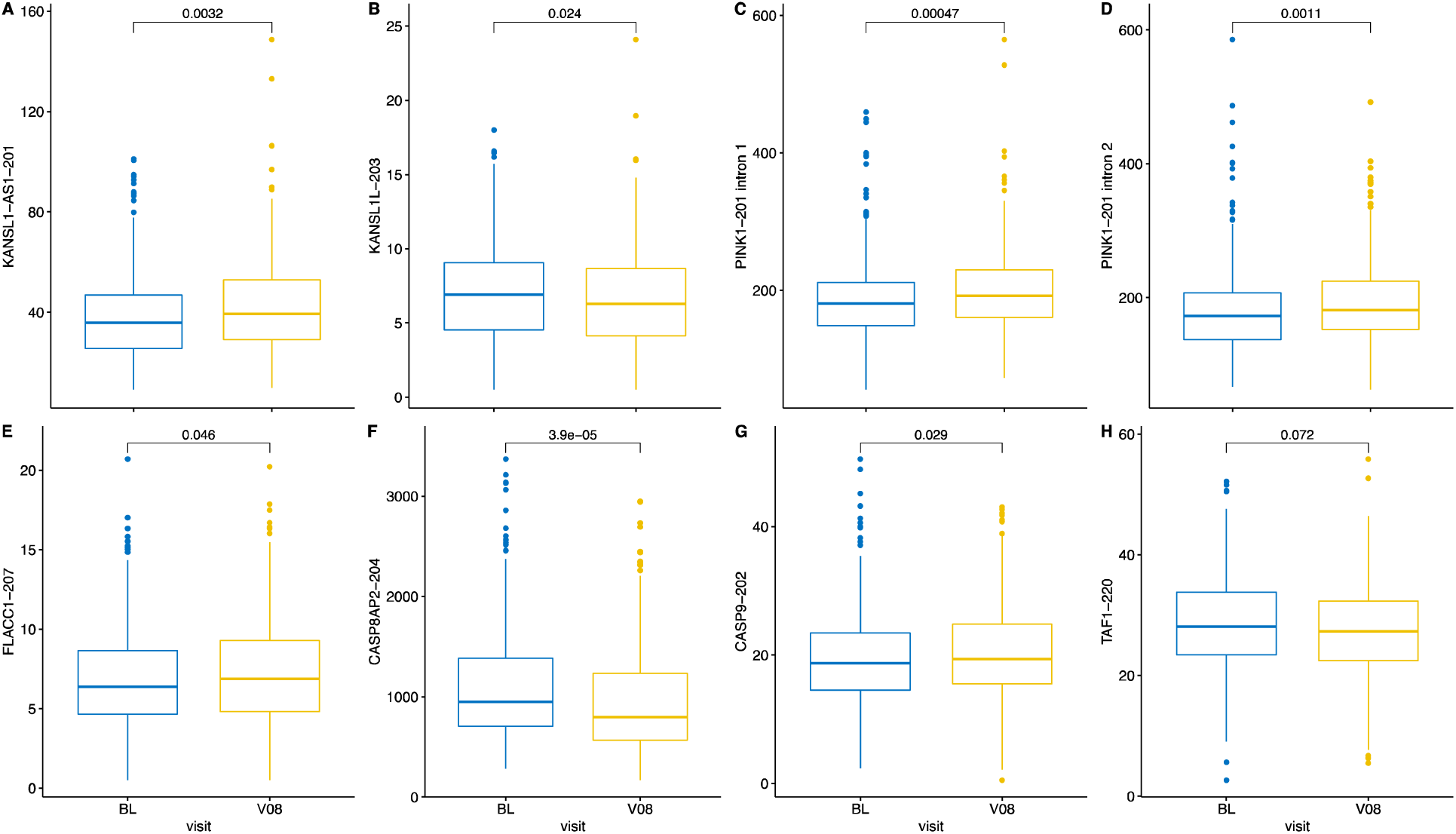
Longitudinal differential expression (normalised counts) of the intronic transcripts in PD patients, BL compared to the V08 visit. The panels A, B, E, F, G, H reflect changes in the introns for different transcripts. Panels C and D illustrate changes for PINK1-201 transcripts, introns 1 and 2.

### Baseline differences

We analysed PD versus CO at different timepoints to have two separate cross-sectional snapshots of transcriptional changes. At the time of diagnosis, PD patients had 836 introns differentially expressed compared to the controls (Supplementary Table 5). Only one exon was differentially expressed in the CO demonstrating again that the enhanced intronic expression is a very specific feature of PD pathogenesis (Supplementary Table 6). Many of the genes identified with differentially expressed introns in PD cohort were involved in signalling pathways which are predicted to impact on progression such as *SNCA, GOLGA5* and *DNAJC19* genes (Figure 4). *GOLGA5* is involved in the Golgi membrane and synaptic vesicle trafficking and docking ^21^. *DNAJC19* or *TIMM14* is a chaperone that functions as an inner mitochondrial membrane co-chaperone and is responsible for mitochondrial protein import, a process that is involved in the pathogenesis of PD among others what other? Neurodegeneration in general/cancer? ^22-24^. The gene *WDFY3* showed eight introns to be differentially upregulated in PD patients (Figure 5). This gene is responsible for mitochondrial quality control and is associated with multiple severe neurological pathologies as it is involved in brain energetics and mitophagy ^25^. We also identified genes with differential intronic upregulation related to the immune response or to the age at onset of PD, such as *PLXNC1, TNFAIP6* and *PRRG4* (Figure 6). These changes were found at the time of diagnosis and may indicate already existing active transcriptional changes from the moment of clinical presentation of PD.

**Figure 4.**
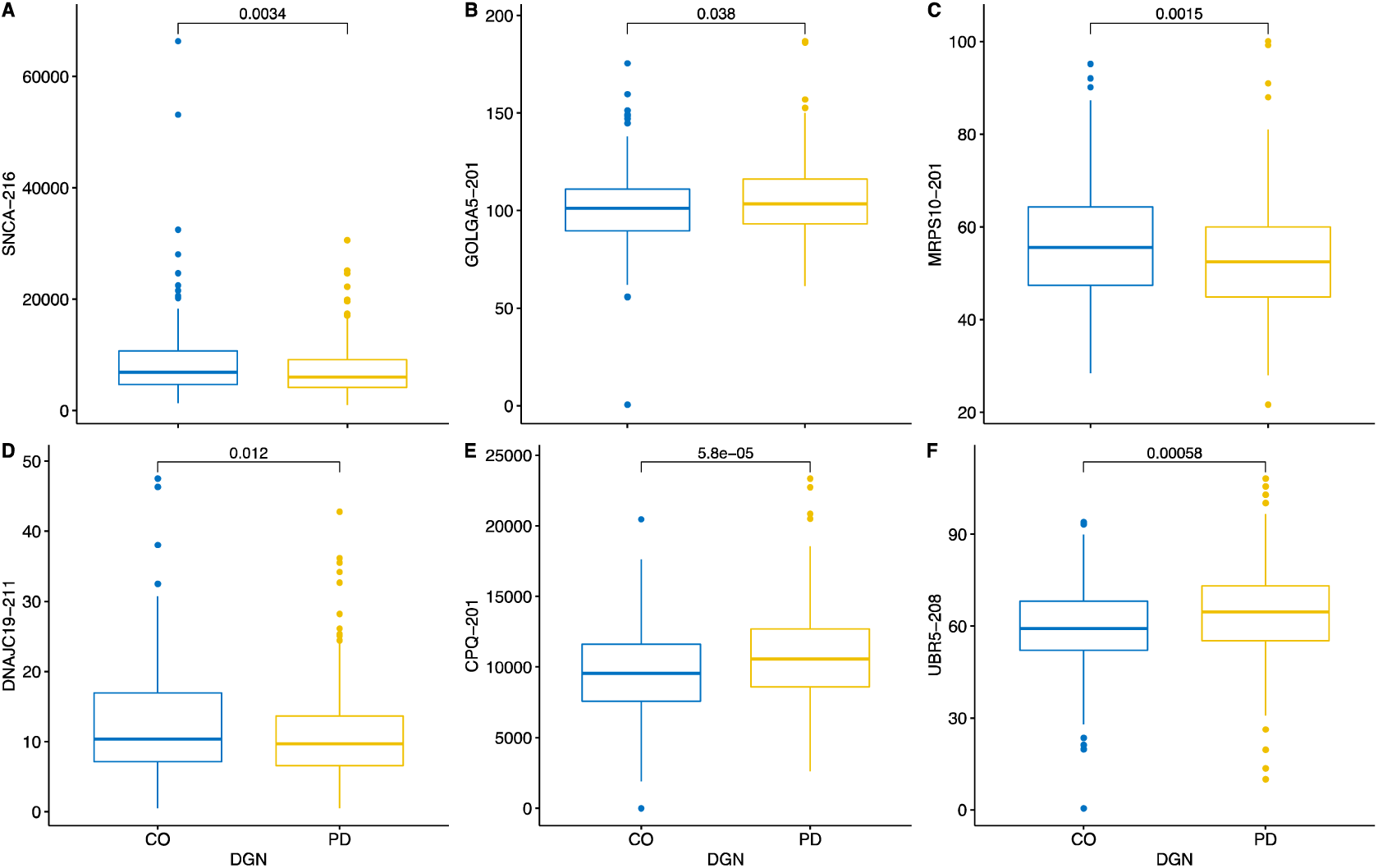
Cross-sectional analysis of the intronic transcripts (normalised counts) at the baseline visit (BL), at the time of enrolment of the subjects. PD patients had decreased expression of the SNCA-216, DNAJC19-211 and MRPS10-201 transcripts. The transcripts GOLGA5-201, CPQ-201 and UBR5-208 were upregulated.

**Figure 5.**
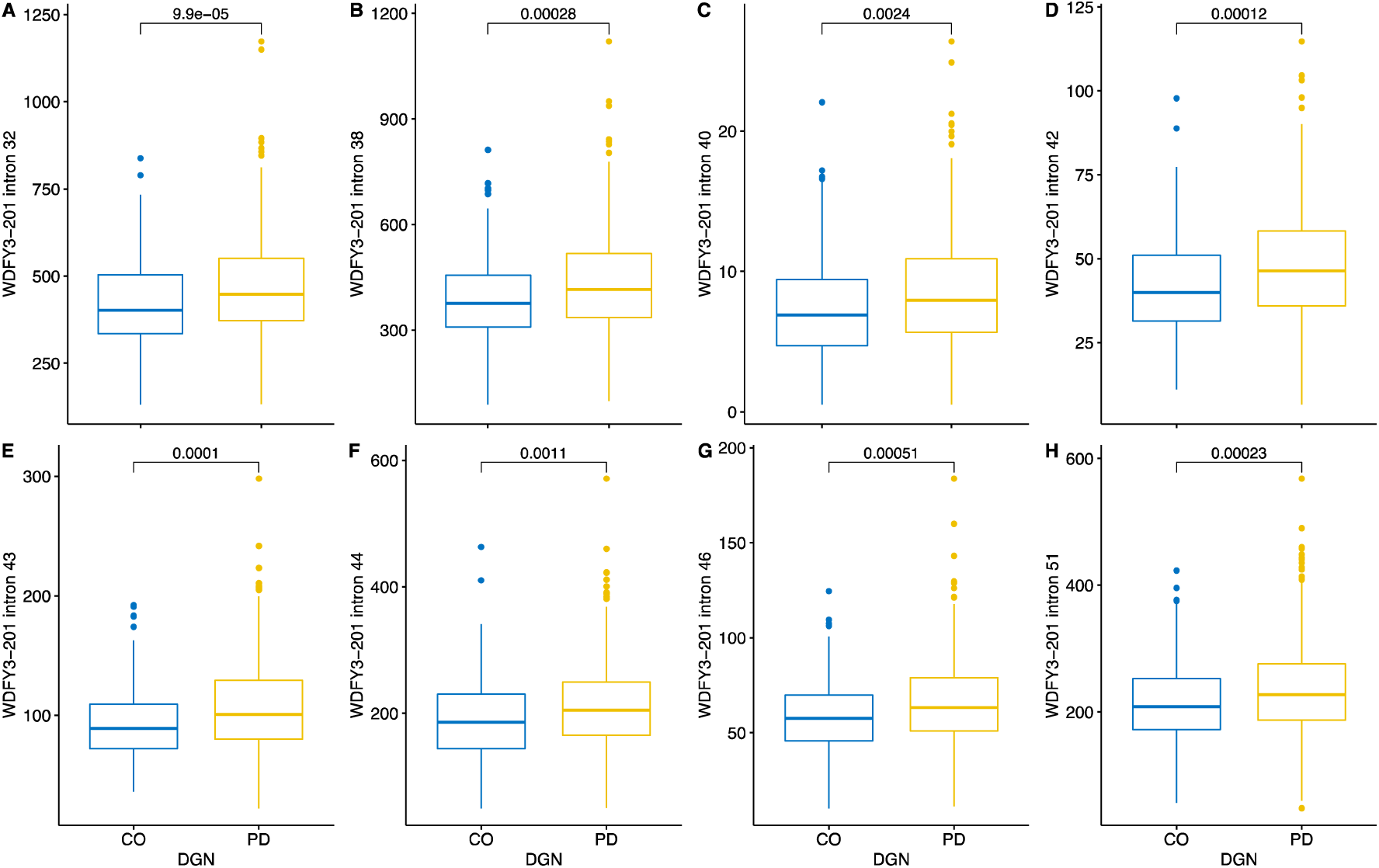
Cross-sectional analysis of the intronic transcripts (normalised counts) at the baseline visit (BL), at the time of enrolment of the subjects. PD patients had increase expression of 8 introns of the WDFY3 gene.

**Figure 6.**
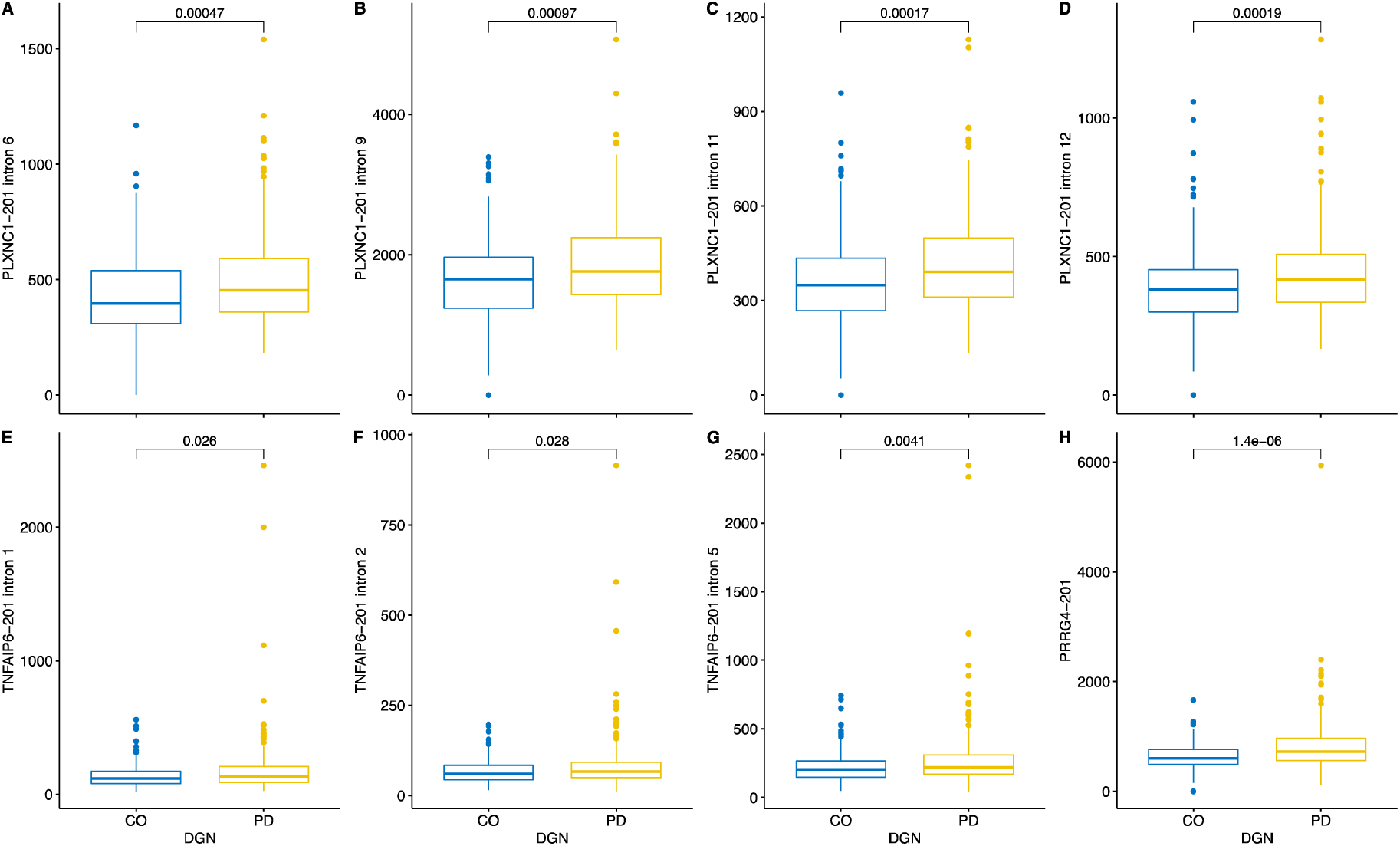
Cross-sectional analysis of the intronic transcripts (normalised counts) at the baseline visit (BL), at the time of enrolment of the subjects. Different introns of the transcripts related to the immune response were up regulated in PD patients.

### Differences at the three-year follow up visit (V08)

While it is important to have a cross sectional snapshot at the time of diagnosis, longitudinal study design allow exploration of time-dependent alterations of the transcriptome. Three years after diagnosis, PD patients had 2,184 intronic transcripts (transcripts containing introns) differentially expressed (Supplementary Table 7). At the same time only 17 differentially expressed intronic transcripts were from exon sequences indicating again very high specificity toward intronic transcription (Supplementary Table 8). From all the detected 2,184 differential intronic transcripts, 329 were identical to the intronic transcripts at the BL timepoint demonstrating major changes in nascent transcriptional changes in the PD group. We identified longitudinal changes in the expression of *GBA-204* and *GBA-206* isoforms, *LRRK2* and *PINK1*; genes that are associated with the pathogenesis of PD from both genomic or functional studies (Figure 7). Only one transcript, *RP11-403I13.4-002*, was identical in the exonic and intronic analysis of V08 timepoint in the PD group.

**Figure 7.**
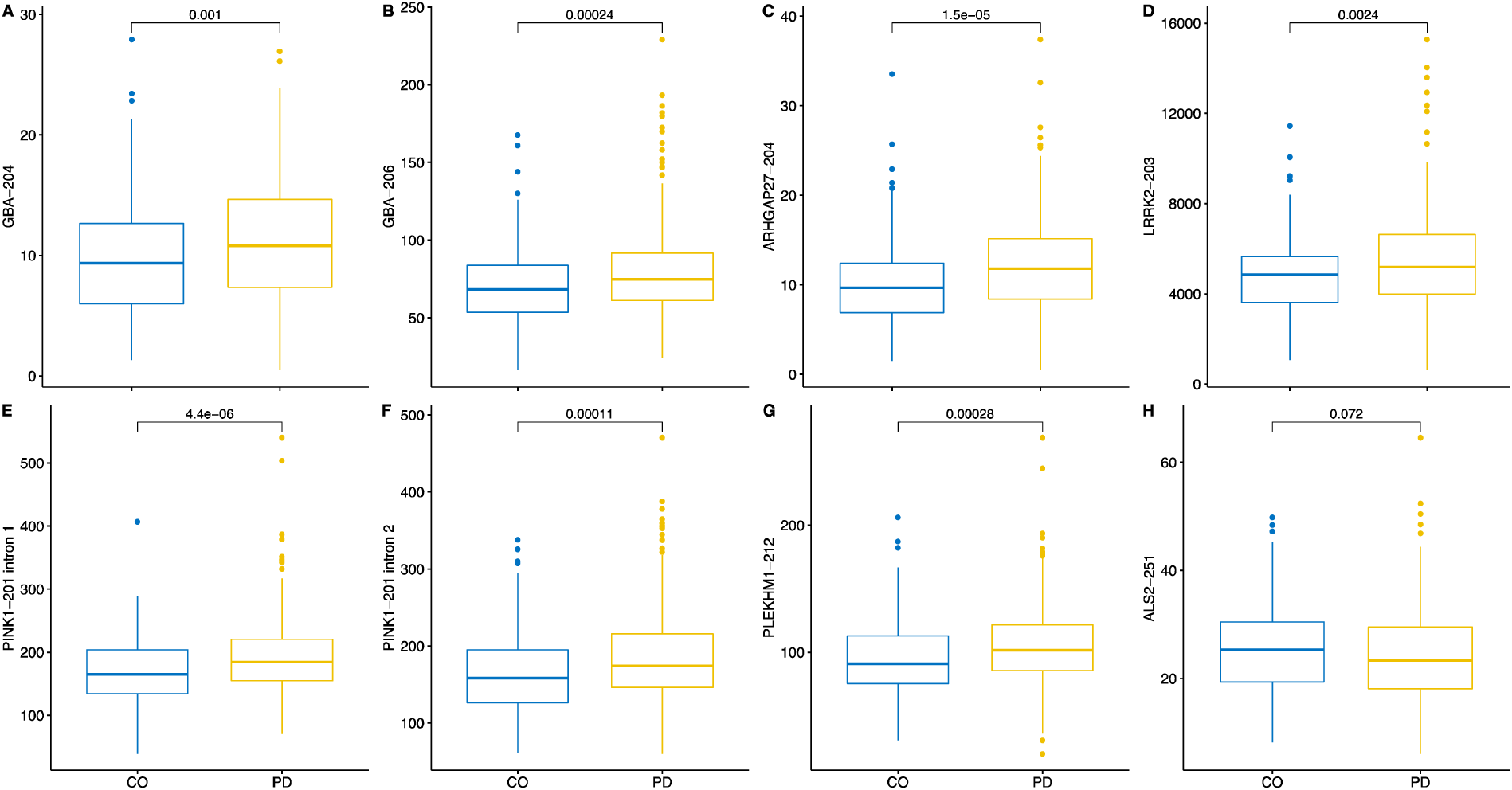
Cross-sectional analysis of the intronic transcripts (normalised counts) at the V08 visit, three years after enrolment of the subjects. Two different transcripts of GBA, LRRK2-203, ARHGAP27-204 and PLEKHM1-212 were upregulated in PD patients. Intorns1 and 2 of the PINK1-201 transcript were also upregulated. ALS2-251 transcript was downregulated in PD patients.

Taken together, transcriptome analysis of introns and exons indicated specific and overwhelming changes in intronic transcription in PD patients compared to CO subjects, these differences were evident at the time of diagnosis and escalated during the three-year progression of the disease.

### Pathway analysis of longitudinal changes

We performed functional pathway analysis to identify the enriched pathways linked to the activated intronic transcription and to identify the common theme of these activated transcripts. The most observed number of changes occurred in the intronic expression over time in the PD cohort, therefore we used only PD data for pathway analysis and compared the intronic transcriptome profiles between the visits BL and V08. Although the analysis was performed on blood RNA-Seq data, reactome enrichment analysis identified that nervous system related themes were enriched in the differential profile of intronic transcription (Figure 8, Table 3). The pathways we identified contained neuronal system, protein-protein interactions at synapses, transmission across chemical synapses and muscle contraction related pathways (Table 2). The activation of neuronal pathways is illustrated in Figure 8 as a bar plot and a dot plot showing the number of genes identified and gene ratio. To illustrate this enrichment further, we used heat and tree plots (Figure 9) showing again the large number of genes mapping to the pathways. This indicated that the longitudinal changes in active transcription in the blood may reflect changes in the nervous system although the mechanism for such mimicking is unclear but could for instance reflect immune parameters. However, the data does indicate that these pathways found in the longitudinal changes reflected PD progression. This is consistent with longitudinal comparison of PD and CO groups separately in which very limited intronic transcriptional alterations in the longitudinal CO group compared to PD group were found.

**Table 3.** Reactome pathway over-representation analysis. Longitudinal changes of the intronic transcription in PD patients were associated with the biological themes related to the nervous system.

| Description | Gene Ratio | P-value | P-adjusted |
| --- | --- | --- | --- |
| Extracellular matrix organization | 80/1505 | $2.6 \times 10^{-9}$ | $3.1 \times 10^{-6}$ |
| Neuronal System | 100/1505 | $4.3 \times 10^{-9}$ | $3.1 \times 10^{-6}$ |
| Protein-protein interactions at synapses | 30/1505 | $6.7 \times 10^{-7}$ | 0.0003 |
| Muscle contraction | 50/1505 | $1.0 \times 10^{-5}$ | 0.003 |
| Striated Muscle Contraction | 14/1505 | 0.0001 | 0.02 |
| Transmission across Chemical Synapses | 59/1505 | 0.0002 | 0.02 |
| Neurexins and neuroligins | 18/1505 | 0.0003 | 0.02 |

**Figure 8.**
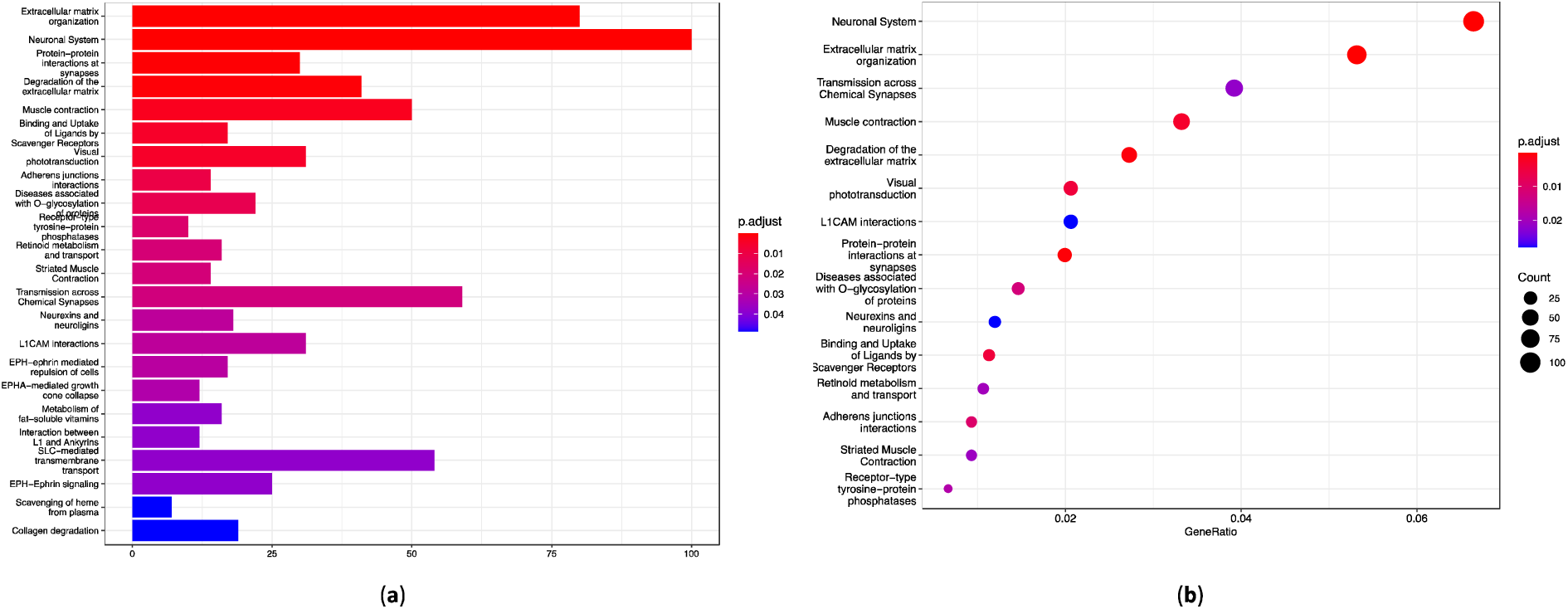
Reactome pathway analysis indicates longitudinal enrichment of the neuronal themes in the blood transcriptome of the PD patients. (**a**) Bar plot of all statistically significant enriched pathways indicates the different number of genes for each pathway. (**b**) Dot plot of the enriched pathways shows the relation between the gene count, gene ratio and adjusted p-values.

**Figure 9.**
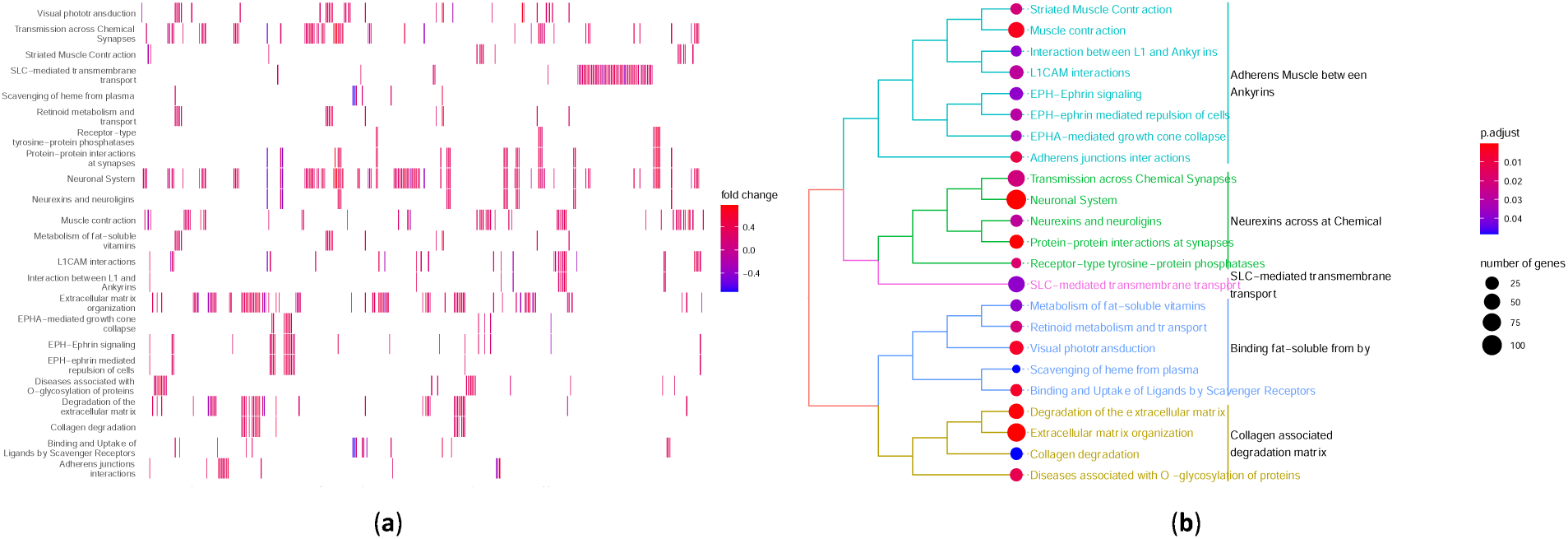
Reactome pathway analysis identified several neuronal pathways enriched longitudinally in PD blood transcriptome. (**a**) Heat plot showing individual genes and their mapping to pathways; (**b**) Tree plot illustrates clustering of the pathways, number of matched genes and adjusted p-values.

KEGG enrichment analysis was performed to validate our previous pathway analysis and to identify potential PD specific pathways (Figures 10 and 11). Indeed, when using KEGG annotation, the most significantly changed pathways were ubiquitin mediated proteolysis, protein processing in endoplasmic reticulum, mitophagy and autophagy. The activation of protein processing in the endoplasmic reticulum (Figure 10) and the activation of mitophagy pathway (Figure 11) during the time course of PD supports the causal relationship between the transcripts and disease progression. All these pathways are directly involved in the pathogenesis of PD.

**Figure 10.**
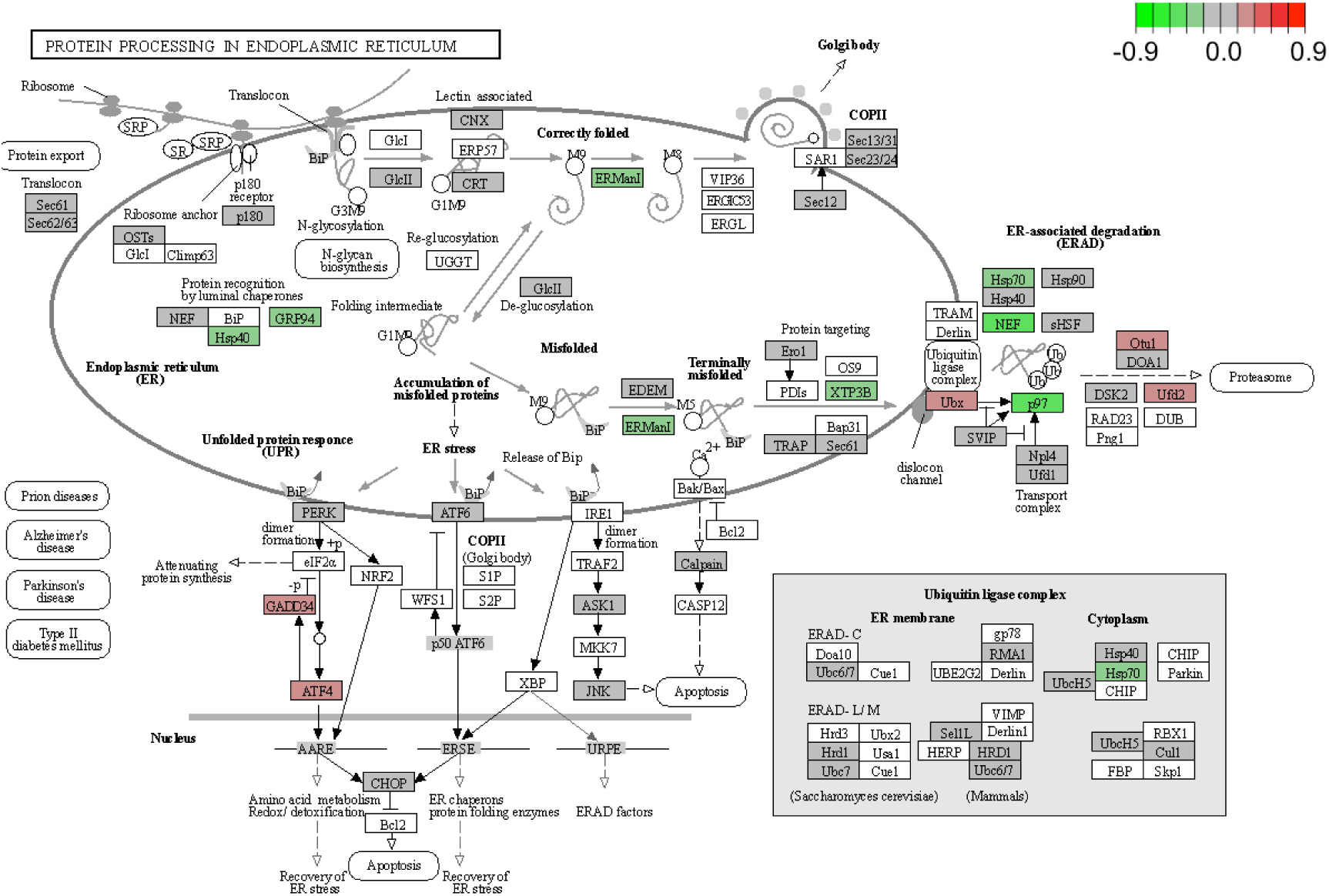
KEGG pathway of the protein processing in the endoplasmic reticulum and the mapped genes from the intronic transcriptome. Mapped genes are in red, green, or grey colors.

**Figure 11.**
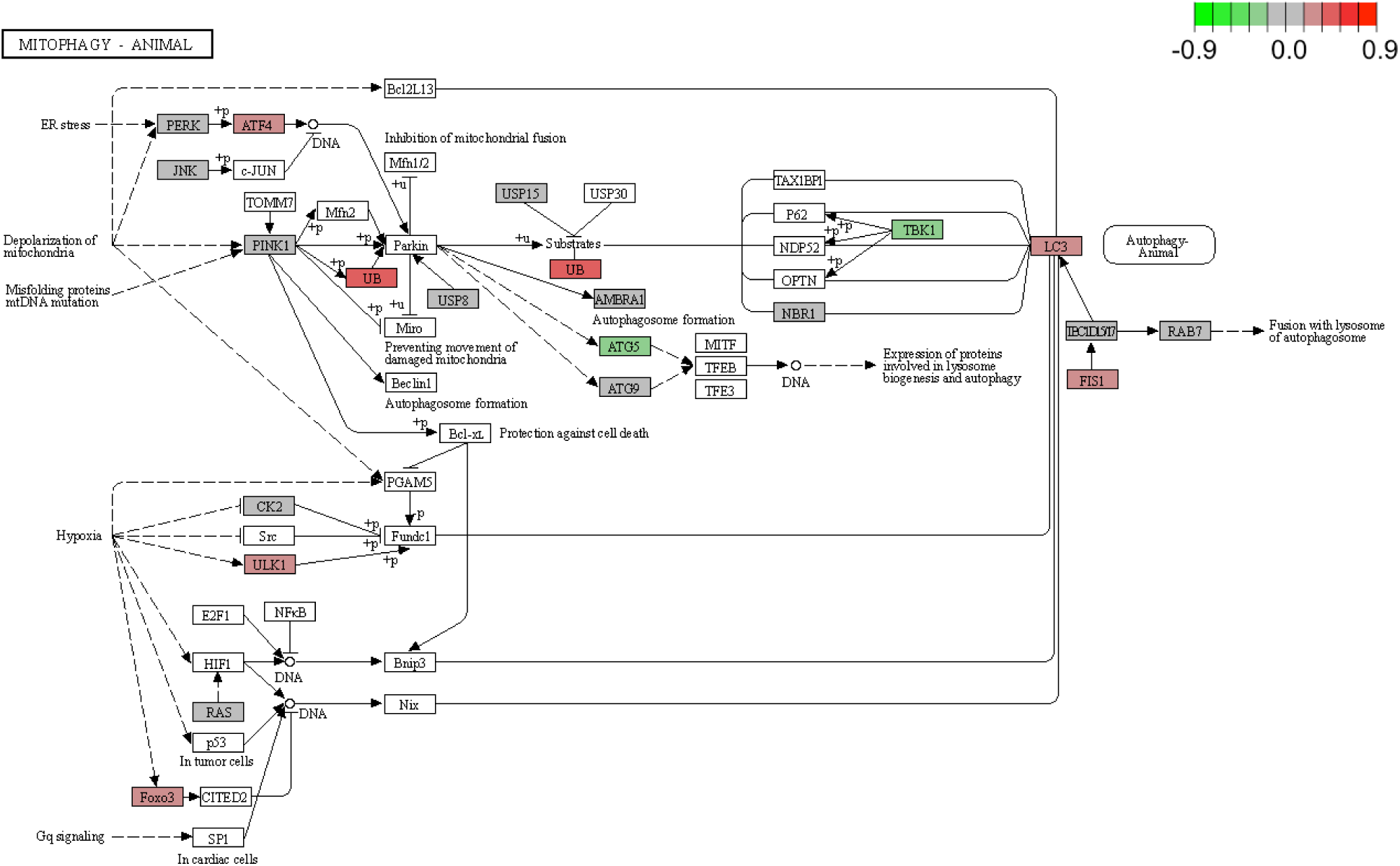
KEGG mitophagy pathway. Longitudinal blood intronic transcriptome profile of Parkinson disease mapped significantly to the mitophagy pathway. Mapped genes are in red, green, or grey colours.

Whilst we analyzed the blood whole transcriptome data, using the intronic reads helped to identify genes with active transcriptional change reflecting the dynamic alterations in the transcriptional balance of the cells both in PD compared to CO and in the progression of PD. We were able to identify within these global changes PD specific pathogenetic networks that have been previously identified to be involved in PD using molecular models and genetic analysis. These results demonstrate that blood transcriptome can reflect nascent transcription specific for the disease condition and indeed potentially progression of the disease.

Taken together, analysis of the intronic reads in the whole genome transcriptome study allows to detect active nascent transcription and allow for more detailed information compared to the exon centric steady-state analysis of transcription alone. We demonstrate that analysis of intronic expression detected actively transcribed transcripts that we were able to map onto functionally and clinically relevant signaling networks that may give us not only additional biomarkers but also new avenues for therapeutic intervention in PD progression.

## Discussion

Whole transcriptome analysis is typically based on gene or exon annotation, quite seldom transcript-based annotation is used ^26, 27^. Gene-based annotation is an amalgamated approach where the reads mapping to different exons and transcripts will be merged under a single functional identifier, the gene ^28^. This approach is the most widely used and therefore most of the whole transcriptome studies provide this aggregated information. This approach leads to the loss of power to detect precise and detailed changes in transcription leading to the loss of the sensitivity in the analysis. The avoidance of transcript-based annotation is understandable as the bioinformatics tools are limited to accurately call transcripts from short read sequence data, the data which is most commonly available to date. Only recently Kallisto and Salmon were developed to provide quasi-mapping approach and accurate transcript calling to overcome the issue with transcript detection ^29, 30^. Nevertheless, using these tools nascent transcription cannot be distinguished from steady state transcription.

Intronic mapping was suggested to be an alternative to identify the transcripts that were recently transcribed ^2^. This approach, based on the co-transcriptional splicing and detection of introns, indicates that some transcripts still have introns included in the RNA-Seq data. Detecting this event in the transcriptional process gives important additional power to measure the genes that are actively and newly transcribed compared to the steady state stable transcription in the background. The analysis of intronic transcripts has successfully been applied to human basal ganglia data with clear evidence for the reproducibility of the intronic eQTLs (i-eQTLs) and their utility to analyze the rate of transcription ^15^. Interestingly, in that paper the authors also identified highly specific enrichment of disease-specific transcription suggesting the suitability of the intron based transcriptional analysis to distinguish steps in the pathogenic process.

In our analysis of intronic expression from the PPMI cohort we have identified longitudinal changes in intronic transcription in PD patients. The PD changes are specific for the disease as only minor differences were found in the analysis of the matched controls. Indeed, functional annotation of the intronic transcripts, although only blood RNA-Seq was analyzed, revealed activation of signaling pathways known to involved to the pathophysiology of PD. These can be utilized as biomarkers of disease and its progression, however their functional significance awaits further analysis such as using patient derived cell lines. Interestingly although the analysis used blood transcriptomic data the identified differences in many cases reflected changes in the pathways related to neurodegenerative processes which might be predicted to occur in the CNS.

Of the many different transcripts identified in the present study there are several involved in mitochondrial function and proteostasis consistent with pathways involved in the pathophysiology and progression of PD. The gene *DNAJC19* or *TIMM14* encodes for an inner mitochondrial membrane translocase that imports proteins into the mitochondria and is directly involved in the pathogenesis of neurodegenerative diseases ^22-24^. Products of TIMM genes interact with translocases of the outer mitochondrial membrane (TOMM) to form a transport pathway for the nuclear encoded precursor proteins ^22^. In a previous study we identified TOMM20 to be involved in the neurodegeneration caused by the downregulation of *WFS1* gene leading to mitochondrial damage ^31^. In addition, in the present study we identified eight introns of the *WDFY3* gene to be differentially upregulated in PD patients. WDFY3 functions as a conductor for aggregate clearance by autophagy and colocalises with the aggregated proteins ^32^. WDFY3 is associated with multiple severe neurological pathologies by regulating brain bioenergetics, autophagy and mitophagy ^25, 33-35^. The findings from single transcripts and intronic changes were further confirmed by functional annotation of differential expression and identification of the activation of autophagy and mitophagy in PD patients. All the above changes in intronic transcription were evident at the time of diagnosis indicating concomitant transcriptional changes with the clinical presentation of the PD.

Comparison of the intronic and exonic signals has recently been systemically analyzed and the reflection of the nascent transcription by the intronic reads confirmed ^2^. Several experimental studies indicate that intronic transcription is a reliable proxy to measure nascent transcription ^7^. Moreover, comparing exonic reads to intronic signals helps to differentiate transcriptional changes from post-transcriptional changes. Therefore, the differences we have identified are caused by changes in active transcription and are not due to changes related to the steady-state processes.

The main limitation of our study is that it is based on short read sequencing data and the power to detect transcripts could be quite limited. However, the separation of individual genomic elements, introns and exons, helps to overcome this issue as the mapping reliability to individual introns and exons is very high. Another limitation of the study is that it is descriptive in the nature and does not provide hard experimental evidence for pathway modulation. The longitudinal design of the study helps somewhat overcome this issue and provides time-dependent changes that could mean a causative relation to the detected changes. The repeated sampling of the cohort makes it possible that the changes we describe here are significant biomarkers for PD progression as we can minimize the effect of the biological variability. Our work gives a comprehensive overview of the time-dependent transcriptional changes in this large PD cohort followed up for many years. The last limitation is more general and is related to almost all genomic studies. Focusing only on the exon-based gene-centered annotation as only “functional genome” ignores the other layers, such as different types of RNAs and introns. A good example of this would be X-linked dystonia-parkinsonism in which intron retention is one of the mechanism thought to be involved in disease progression (TAF1 ref to be added). Our data may be a first step more generally to improve our knowledge about the intronic transcription and how these changes can be involved in the pathogenesis of the disease.

## Conclusions

In conclusion, we identified highly specific longitudinal nascent transcriptional profile in the blood of Parkinson patients that possibly reflects the changes caused by the molecular pathological processes of the disease and are relevant to improve our understanding about the progression of the disease.

## Data Availability

All data produced are available online at www.ppmi-info.org.

## Authors’ Contributions

Conceptualization, S.K.; methodology, A.L.P. and S.K.; formal analysis, S.K.; data interpretation, A.L.P., V.J.B., J.P.Q. and S.K.; writing—original draft preparation, S.K.; writing—review and editing, A.L.P., V.J.B., J.P.Q. and S.K.; funding acquisition, A.L.P. and S.K. All authors have read and agreed to the published version of the manuscript.

## Data Availability

Raw data are available from the PPMI website (www.ppmi-info.org/data (accessed on 19 January 2021)).

